# Performance of ChatGPT on USMLE: Potential for AI-Assisted Medical Education Using Large Language Models

**DOI:** 10.1101/2022.12.19.22283643

**Authors:** Tiffany H. Kung, Morgan Cheatham, ChatGPT, Arielle Medenilla, Czarina Sillos, Lorie De Leon, Camille Elepaño, Maria Madriaga, Rimel Aggabao, Giezel Diaz-Candido, James Maningo, Victor Tseng

## Abstract

We evaluated the performance of a large language model called ChatGPT on the United States Medical Licensing Exam (USMLE), which consists of three exams: Step 1, Step 2CK, and Step 3. ChatGPT performed at or near the passing threshold for all three exams without any specialized training or reinforcement. Additionally, ChatGPT demonstrated a high level of concordance and insight in its explanations. These results suggest that large language models may have the potential to assist with medical education, and potentially, clinical decision-making.

## INTRODUCTION

Over the past decade, advances in neural networks, deep learning, and artificial intelligence (AI) have transformed the way we approach a wide range of tasks and industries ranging from manufacturing and finance to consumer products. The ability to build highly accurate classification models rapidly and regardless of input data type (e.g. images, text, audio) has enabled widespread adoption of applications such as automated tagging of objects and users in photographs^1^, near-human level text translation^2^, automated scanning in bank ATMs, and even the generation of image captions^3^.

While these technologies have made significant impacts across many industries, applications in clinical care remain limited. The proliferation of clinical free-text fields combined with a lack of general interoperability between health IT systems contribute to a paucity of structured, machine-readable data required for the development of deep learning algorithms. Even when algorithms applicable to clinical care are developed, their quality tends to be highly variable, with many failing to generalize across settings due to limited technical, statistical, and conceptual reproducibility^4^. As a result, the overwhelming majority of successful healthcare applications currently support back-office functions ranging from payor operations, automated prior authorization processing, and management of supply chains and cybersecurity threats. With rare exceptions – even in medical imaging – there are relatively few applications of AI directly used in widespread clinical care today.

The proper development of clinical AI models^5^ requires significant time, resources, and more importantly, highly domain and problem-specific training data, all of which are in short supply in the world of healthcare. One of the key developments that enabled image-based AI in clinical imaging has been the ability of large general domain models to perform as well as, or even outperform, domain-specific models. This development has catalyzed significant AI activity in medical imaging, where otherwise it would be challenging to obtain sufficient annotated clinical images. Indeed today, Inception-V3 serves as the basic foundation of many of the top medical imaging models currently published, ranging from ophthalmology^5,6^, pathology^7^, to dermatology^8^.

In the past three weeks, a new AI model called ChatGPT captured significant attention due to its ability to perform a diverse array of natural language tasks^9^. ChatGPT is a general Large Language Model (LLM) developed recently by OpenAI. While the previous class of AI models have primarily been Deep Learning (DL) models, which are designed to learn and recognize patterns in data, LLMs are a new type of AI algorithm trained to predict the likelihood of a given sequence of words based on the context of the words that come before it. Thus, if LLMs are trained on sufficiently large amounts of text data, they are capable of generating novel sequences of words never observed previously by the model, but that represent plausible sequences based on natural human language. ChatGPT is powered by GPT3.5, an LLM trained on the OpenAI 175B parameter foundation model and a large corpus of text data from the Internet via reinforcement and supervised learning methods. Anecdotal usage indicates that ChatGPT exhibits evidence of deductive reasoning and chain of thought, as well as long-term dependency skills.

In this study, we evaluate the performance of ChatGPT, a non-domain specific LLM, on its ability to perform clinical reasoning by testing its performance on questions from the United States Medical Licensing Examination (USMLE). The USMLE is a high-stakes, comprehensive three-step standardized testing program covering all topics in physicians’ fund of knowledge, spanning basic science, clinical reasoning, medical management, and bioethics. The difficulty and complexity of questions is highly standardized and regulated, making it an ideal input substrate for AI testing. The examination is well-established, showing remarkably stable raw scores and psychometric properties over the previous ten years^10^. The Step 1 exam is typically taken by medical students who have completed two years of didactic and problem-based learning and focuses on basic science, pharmacology, and pathophysiology; medical students often spend approximately 300-400 hours of dedicated study time in preparation for this exam^11^. The Step 2CK exam is usually taken by fourth-year medical students who have additionally completed 1.5 to 2 years of clinical rotations; it emphasizes clinical reasoning, medical management, and bioethics. The Step 3 exam is taken by physicians who generally have completed at least a 0.5 to 1 year of postgraduate medical education.

USMLE questions are textually and conceptually dense; text vignettes contain multimodal clinical data (i.e., history, physical examination, laboratory values, and study results) often used to generate ambiguous scenarios with closely-related differential diagnoses. Due to its linguistic and conceptual richness, we reasoned that the USMLE would serve as an excellent challenge for ChatGPT.

Our work aims to provide both qualitative and quantitative feedback on the performance of ChatGPT and assess its potential for use in healthcare.

## METHODS

### Artificial Intelligence

ChatGPT (OpenAI; San Francisco, CA), is a large language model that uses self-attention mechanisms and a large amount of training data to generate natural language responses to text input in a conversational context. It is particularly effective at handling long-range dependencies and generating coherent and contextually appropriate responses. ChatGPT is a server-contained language model that is unable to browse or perform internet searches. Therefore, all responses are generated *in situ*, based on the abstract relationship between words (“tokens”) in the neural network. This contrasts to other chatbots or conversational systems that are permitted to access external sources of information (e.g. performing online searches or accessing databases) in order to provide directed responses to user queries.

### Input Source

376 publicly-available test questions were obtained from the June 2022 sample exam release on the official USMLE website. Random spot checking was performed to ensure that none of the answers, explanations, or related content were indexed on Google prior to January 1, 2022, representing the last date accessible to the ChatGPT training dataset. All sample test questions were screened, and questions containing visual assets such as clinical images, medical photography, and graphs were removed. After filtering, 305 USMLE items (Step 1: 93, Step 2CK: 99, Step 3: 113) were advanced to encoding.

### Encoding

Questions were formatted into three variants and input into ChatGPT in the following sequence:

1. Open-ended (OE) format: Created by removing all answer choices, adding a variable lead-in interrogative phrase. This format simulates free input and a natural user query pattern.
2. Multiple choice single answer without forced justification (MC-NJ): Created by reproducing the original USMLE question verbatim.
3. Multiple choice single answer with forced justification (MC-J): Created by adding a variable lead-in imperative or interrogative phrase mandating ChatGPT to provide a rationale for each answer choice.

Encoders employed deliberate variation in the lead-in prompts to avoid systematic errors that could be caused by stereotyped wording. To reduce memory retention bias, a new chat session was started in ChatGPT for each entry. Post-hoc analyses were performed to exclude systematic variation by encoder (data not shown).

### Adjudication

AI outputs were independently scored for Accuracy, Concordance, and Insight by two physician adjudicators using the rubric provided in **Supplemental Table 1**. To minimize within-item anchoring bias, adjudicators scored Accuracy for all items, followed by Concordance for all items, followed by Insight for all items. To minimize interrater cross-contamination, Physician 1 adjudicated Accuracy while Physician 2 adjudicated Concordance, and so forth. If consensus was not achieved for all three domains, the item was referred to a final physician adjudicator. Only 11 items (3.6% of the dataset) required arbitration.

A schematic overview of the experimental protocol is provided in **Figure 1**.

**Figure 1.**
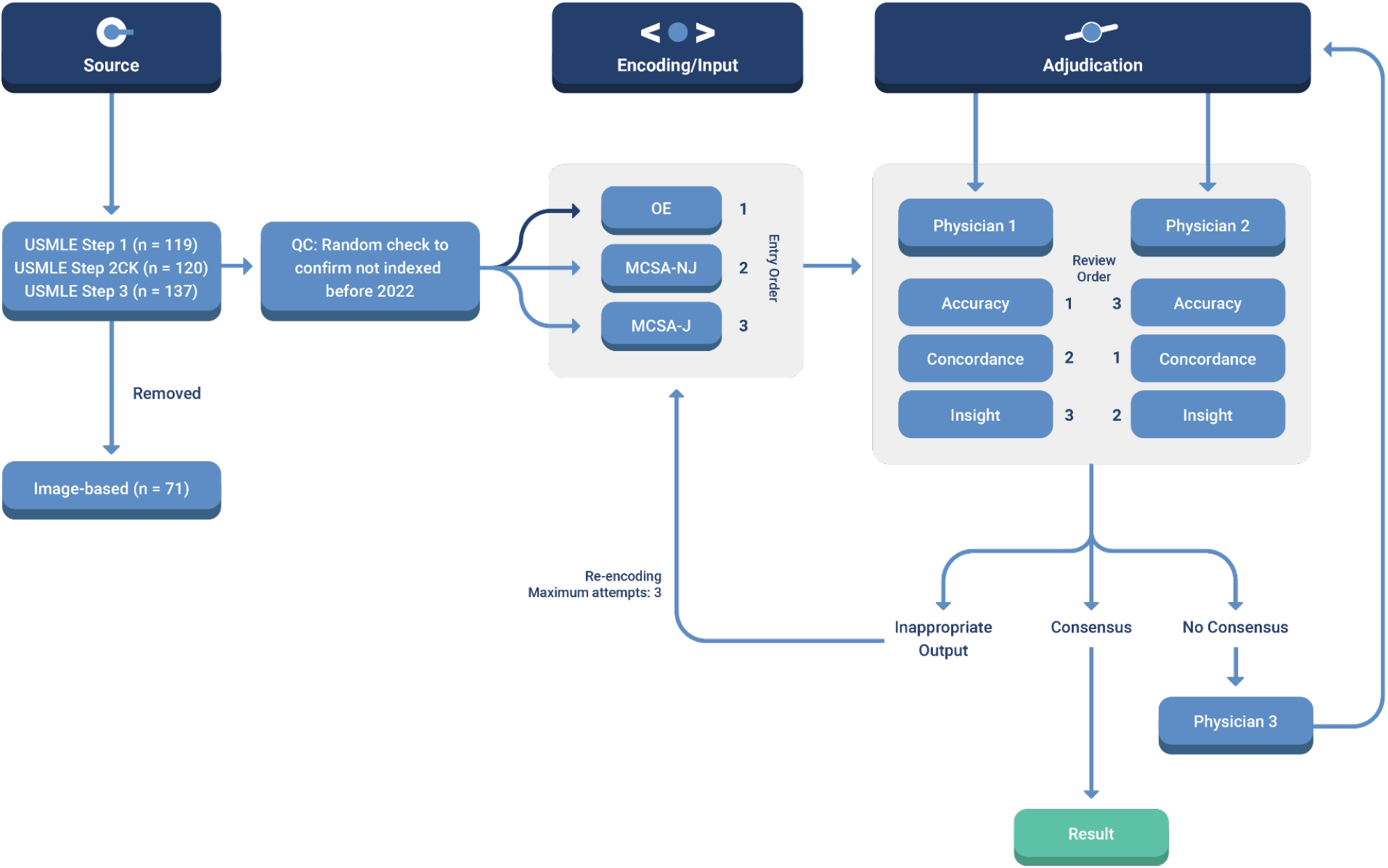
Schematic of workflow for sourcing, encoding, and adjudicating results. Abbreviations: **QC** = quality control; **MCSA-NJ** = multiple choice single answer without forced justification; **MCSA-J** = multiple choice single answer with forced justification; **OE** = open-ended question format

## DATA AVAILABILITY

The data analyzed in this study were obtained from USMLE sample questions sets which are publicly available. The question index, raw inputs, and raw AI outputs are available in the **Online Data Supplement**. Inquiries and requests for additional dataset items and adjudication results can be provided upon reasonable request by contacting Victor Tseng, MD.

## RESULTS

### ChatGPT yields moderate accuracy approaching passing performance on USMLE

Exam items were first encoded as open-ended questions with variable lead-in prompts. This input format simulates a free natural user query pattern. With indeterminate responses censored/included, ChatGPT accuracy for USMLE Steps 1, 2CK, and 3 was 68.0%/42.9%, 58.3%/51.4%, and 62.4%/55.7%, respectively (**Figure 2A**).

**Figure 2.**
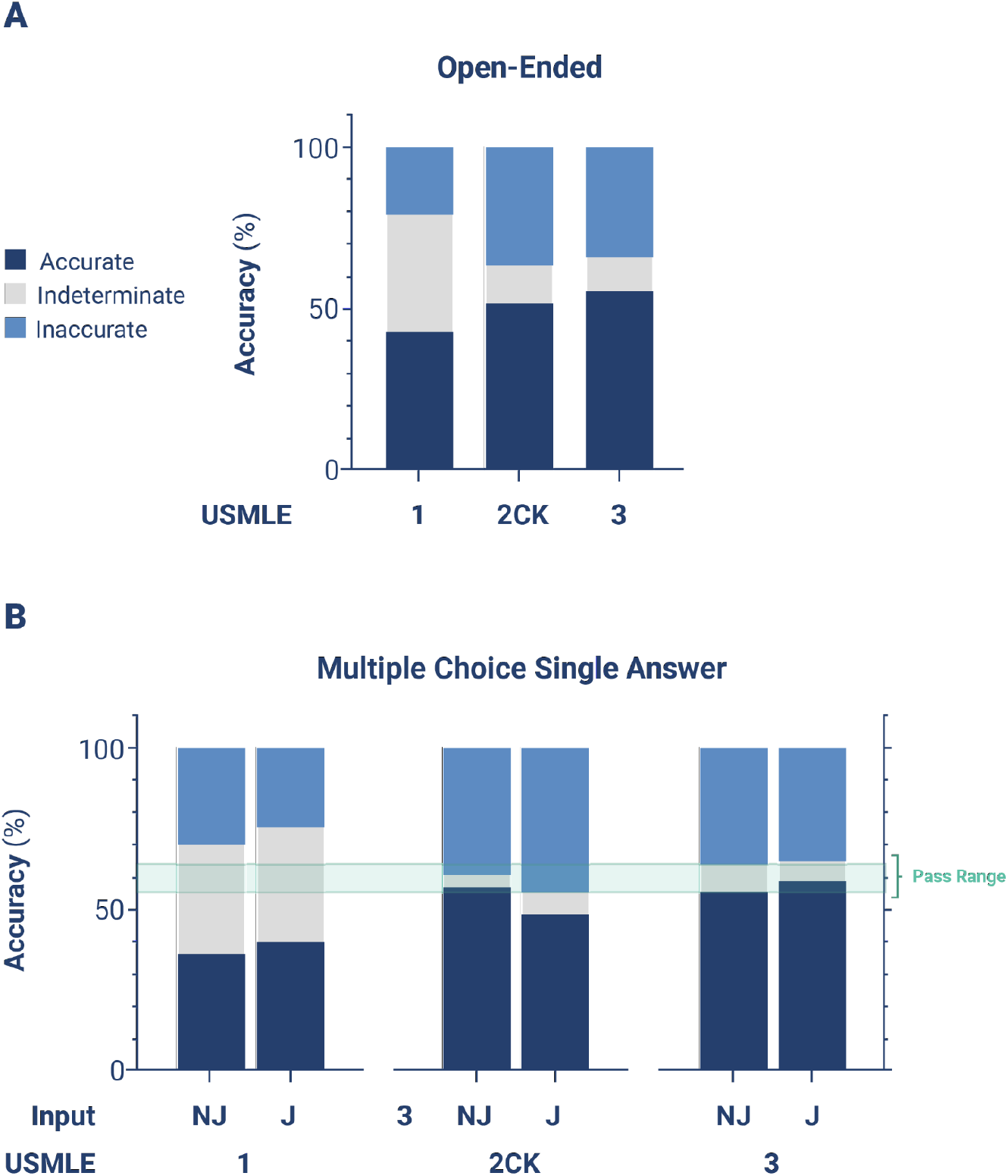
Accuracy of ChatGPT on USMLE. For USMLE Steps 1, 2CK, and 3, AI outputs were adjudicated to be accurate, inaccurate, or indeterminate based on the ACI scoring system provided in Supplemental Table 1. **A:** Accuracy distribution for inputs encoded as open-ended questions **B:** Accuracy distribution for inputs encoded as multiple choice single answer without (MC-NJ) or with forced justification (MC-J)

Next, exam items were encoded as multiple choice single answer questions with no forced justification (MC-NJ). This input is the verbatim question format presented to test-takers. With indeterminate responses censored/included, ChatGPT accuracy for USMLE Steps 1, 2CK, and 3 was 55.1%/36.1%, 59.1%/56.9%, and 60.9%/54.9%, respectively.

Finally, items were encoded as multiple choice single answer questions with forced justification of positive and negative selections (MC-J). This input format simulates insight-seeking user behavior. With indeterminate responses censored/included, ChatGPT accuracy was 62.3%/ 40.3%, 51.9%/48.6%, and 64.6%/59.8%, respectively (**Figure 2B**).

### ChatGPT demonstrates high internal concordance

Concordance was independently adjudicated by two physician reviewers by inspection of the explanation content. Overall, ChatGPT outputted answers and explanations with 94.6% concordance across all questions. High global concordance was sustained across all exam levels, and across OE, MC-NJ, and MC-J question input formats (**Figure 3A**).

**Figure 3.**
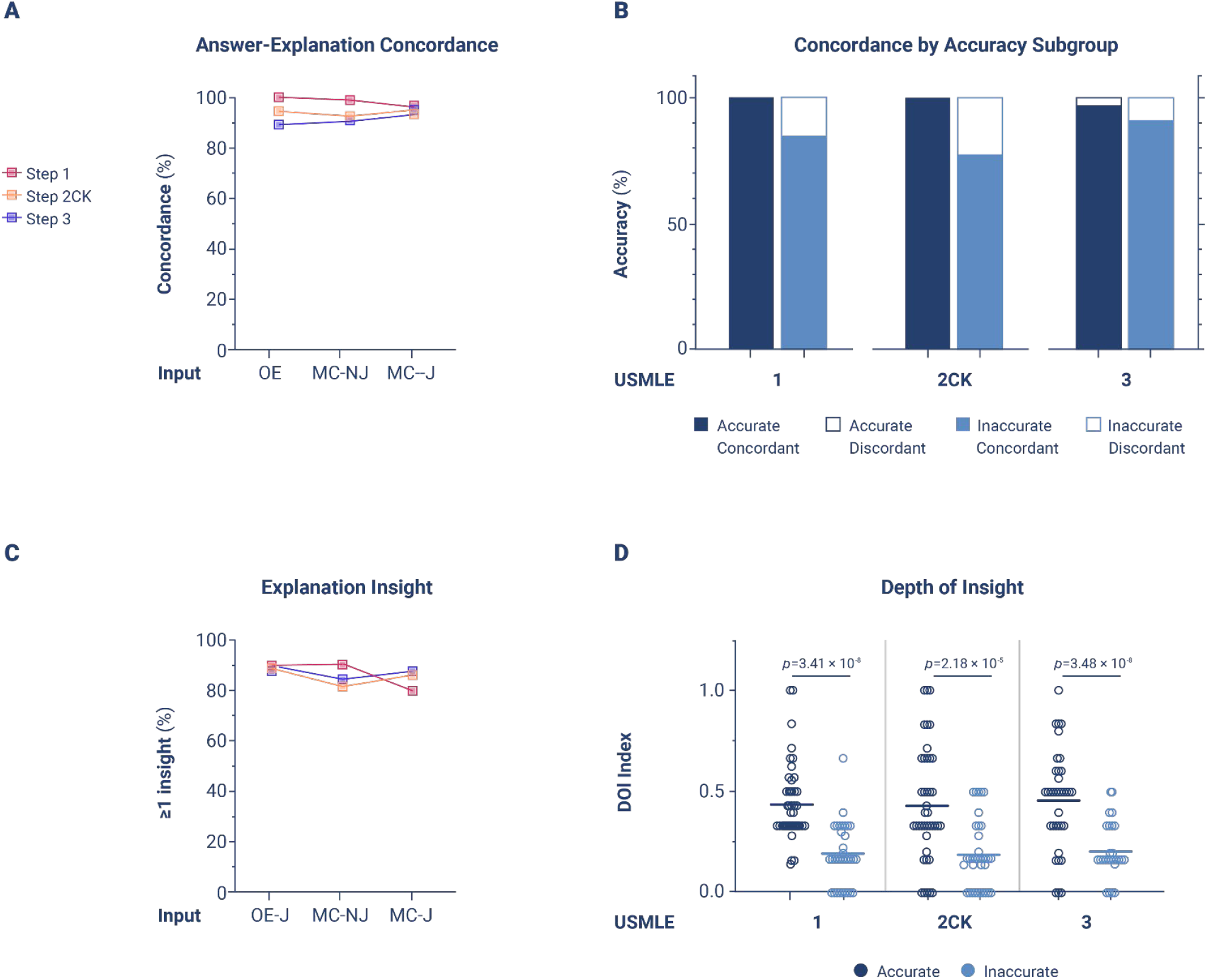
Concordance and insight of ChatGPT on USMLE. For USMLE Steps 1, 2CK, and 3, AI outputs were adjudicated on concordance and density of insight (DOI) based on the ACI scoring system provided in Supplemental Table 1. **A:** Overall concordance across all exam types and question encoding formats **B:** Concordance rates stratified between accurate *vs* inaccurate outputs, across all exam types and question encoding formats. *p* <0.001 for accurate *vs* inaccurate outputs by Fisher exact test **C:** Overall insight prevalence, defined as proportion of outputs with ≥1 insight, across all exams for questions encoded in MC-J format **D:** DOI stratified between accurate *vs* inaccurate outputs, across all exam types for questions encoded in MC-J format. Horizontal line indicates the mean. *p*-value determined by parametric 2-way ANOVA testing with Benjamini-Krieger-Yekutieli (BKY) *post hoc* to control for false discovery rate.

Next, we analyzed the contingency between accuracy and concordance in MC-J responses. ChatGPT was forced to justify its answer choice preference, and to defend its rejection of alternative choices.

Concordance amongst accurate responses was nearly perfect, and significantly greater than amongst inaccurate responses (99.1% vs. 85.1%, *p*<0.001) (**Figure 3B**).

These data indicate that ChatGPT exhibits very high answer-explanation concordance, likely reflecting high internal consistency in its probabilistic language model.

### Generative insight offered by ChatGPT may assist the human learner

Having established the accuracy and concordance of ChatGPT, we next examined its potential to augment human learning in the domain of medical education. AI-generated explanations were independently adjudicated by 2 physician reviewers. Explanation content was examined for significant insights, defined as instances that met the criteria (see **Supplemental Table 1**) of *novelty, nonobviousness*, and *validity*. The perspective of the target test audience was adopted by the adjudicator, as a second-year medical student for Step 1, fourth-year medical student for Step 2CK, and post-graduate year 1 resident for Step 3.

Overall, ChatGPT produced at least one significant insight in 88.9% of all responses. The prevalence of insight was generally consistent between exam type and question input format (**Figure 3C**). In Step 2CK however, insight decreased by 10.3% (*n* = 11 items) between MC-NJ and MC-J formulations. Review of this subset of questions did not reveal a discernible pattern for the paradoxical decrease (see **Supplemental Table 2B**).

To quantify the density of insight (DOI) contained within AI-generated explanations, the number of unique insights was normalized to the number of possible answer choices. This analysis was performed on MC-J entries only. High quality outputs were generally characterized by DOI >0.6 (i.e. unique, novel, nonobvious, and valid insights provided for >3 out of 5 choices); low quality outputs were generally characterized by DOI ≤0.2. The upper limit on DOI is only bounded by the maximum length of text output. Across all exam types, we observed that DOI was significantly higher in questions items answered accurately versus inaccurately (0.458 versus 0.199%, *p* <0.0001) (**Figure 3D**).

These data indicate that a target human learner (e.g., such as a second-year medical student preparing for Step 1), if answering incorrectly, is likely to gain new or remedial insight from the ChatGPT AI output. Conversely, a human learner, if answering correctly, is less likely, but still able to access additional insight.

## DISCUSSION

In this study, we provide new and surprising evidence that ChatGPT is able to perform several intricate tasks relevant to handling complex medical and clinical information. To assess ChatGPT’s capabilities against biomedical and clinical questions of *standardized* complexity and difficulty, we tested its performance characteristics on the United States Medical Licensing Examination (USMLE).

Our findings can be organized into two major themes: (1) the rising accuracy of ChatGPT, which approaches or exceeds the passing threshold for USMLE; and (2) the potential for this AI to generate novel insights that can assist human learners in a medical education setting.

### The rising accuracy of ChatGPT

The most recent iteration of the GPT LLM (GPT3) achieved 46% accuracy with zero prompting^12^, which marginally improved to 50% with further model training. Previous models, merely months prior, performed at 36.7%^13^. In this present study, ChatGPT performed at >50% accuracy across all examinations, exceeding 60% in most analyses. The USMLE pass threshold, while varying by year, is approximately 60%. Therefore, ChatGPT is now comfortably within the passing range. Being the first experiment to reach this benchmark, we believe this is a surprising and impressive result. Moreover, we provided no prompting or training to the AI, minimized grounding bias by expunging the AI session prior to inputting each question variant, and avoided chain-of-thought biasing by requesting forced justification only as the final input. Further model interaction and prompting could often produce more accurate results (data not shown). Given this trajectory, it is likely that AI performance will continue to rise as LLM models continue to mature.

Paradoxically, ChatGPT outperformed PubMedGPT (accuracy 50.8%, unpublished data), a counterpart LLM with similar neural structure, but trained exclusively on biomedical domain literature. We speculate that domain-specific training may have created greater ambivalence in the PubMedGPT model, as it absorbs real-world text from ongoing academic discourse that tends to be inconclusive, contradictory, or highly conservative or noncommittal in its language. A foundation LLM trained on general content, such as ChatGPT, may therefore have an advantage because it is also exposed to broader clinical content, such as patient-facing disease primers and provider-facing drug package inserts, that are more definitive and congruent.

Consistent with the mechanism of generative language models, we observed that the accuracy of ChatGPT was strongly mediated by concordance and insight. High accuracy outputs were characterized by high concordance and high density of insight. Poorer accuracy was characterized by lower concordance and a poverty of insight. Therefore, inaccurate responses were driven primarily by missing information, leading to diminished insight and indecision in the AI, rather than overcommitment to the incorrect answer choice. These findings indicate that model performance could be significantly improved by merging foundation models, such as ChatGPT, with a domain-specific LLM or other model trained on a voluminous and highly validated medical knowledge resources, such as UpToDate, or other ACGME-accredited content.

Interestingly, the accuracy of ChatGPT tended to be lowest for Step 1, followed by Step 2CK, followed by Step 3. This mirrors both the subjective difficulty and objective performance for real-world test takers on Step 1, which is collectively regarded as the most difficult exam of the series. The low accuracy on Step 1 could be explained by an undertrained model on the input side (e.g. underrepresentation of basic science content on the general information space) and/or the human side (e.g. insufficient or invalid human judgment at initial reinforcement stages). This result exposes a key vulnerability in pre-trained LLMs, such as ChatGPT: AI ability becomes yoked to human ability. ChatGPT’s performance on Step 1 is poorer precisely because human users perceive its subject matter (e.g., pathophysiology) as more difficult or opaque.

### The potential for AI-assisted human learning in medical education

We also examined the ability of ChatGPT to assist the human learning process of its target audience (e.g., a second year medical student preparing for USMLE Step 1). As a proxy for the metric of helpfulness, we assessed the concordance and insight offered by the AI explanation outputs. ChatGPT responses were highly concordant, such that a human learner could easily follow the internal language, logic, and directionality of relationships contained within the explanation text (e.g., adrenal *hyper*cortisolism ⥬ *increased* bone osteoclast activity ⥬ *increased* calcium resorption ⥬ *decreased* bone mineral density ⥬ *increased* fracture risk). High internal concordance and low self-contradiction is a proxy of sound clinical reasoning and an important metric of explanation quality. It is reassuring that the directionality of relationships is preserved by the language processing model, where each verbal object is individually lemmatized.

AI-generated responses also offered significant insight, role-modeling a deductive reasoning process valuable to human learners (see **Supplemental Table 2**). At least one significant insight was present in approximately 90% of outputs. ChatGPT therefore possesses the partial ability to teach medicine by surfacing novel and nonobvious concepts that may not be in learners’ sphere of awareness. This qualitative gain provides a basis for future real-world studies on the efficacy of generative AI to augment the human medical education process. For example, longitudinal exam performance can be studied in a quasi-controlled in AI-assisted and unassisted learners. Unit economic analysis may clarify the cost-effectiveness of incremental student performance gain in comparison to existing tools such as virtual tutors and study aids.

Medical education, licensing examinations, and test preparation services form a large industrial complex eclipsing a nine-figure market size annually. While its relevance remains debated, standardized testing has emerged as an important end-target of medical learning. In parallel, of the didactic techniques, a socratic teaching style is favored by medical students^14^. The rate-limiting step for fresh content generation is the human cognitive effort required to craft realistic clinical vignettes that probe “high-yield” concepts in a subtle way, engage critical thinking, and offer pearls of knowledge even if answered incorrectly. Demand for new examination content continues to increase. For a national medical examiner, a single item typically requires 0.1 FTE work effort to produce (NBME, personal communication). Future studies may investigate the ability of generative language AI to offload this human effort by assisting in the question-explanation writing process or, in some cases, writing entire items autonomously.

Finally, the advent of AI in medical education demands an open science research infrastructure to standardize experimental methods, readouts, and benchmarks to describe and quantify human-AI interactions. Multiple dimensions must be covered, including user experience, learning environment, hybridization with other teaching modes, and effect on cognitive bias. In this report, we provide an initial basic protocol for adjudicating AI-generated responses along axes of accuracy, concordance, and insight.

Our study has several important limitations. The relatively small input size restricted the depth and range of analyses. For example, stratifying the output of ChatGPT by subject taxonomy (e.g., pharmacology, bioethics) or competency type (e.g., differential diagnosis, management) may be of great interest to medical educators, and could reveal heterogeneities in performance across language processing for different clinical reasoning tasks. Similarly, a more robust AI failure mode analysis (e.g., language parsing error) may lend insight into the etiology of inaccuracy and discordance. In addition to being laborious, human adjudication is error-prone and subject to greater variability and bias. Future studies will undoubtedly apply unbiased approaches, using quantitative natural language processing and text mining tools such as word network analysis. In addition to increasing validity and accelerating throughput by several orders of magnitude, these methods are likely to better characterize the depth, coherence, and learning value of AI output. Finally, to truly assess the utility of generative language AI for medical education, ChatGPT and related applications must be studied in both controlled and real-world learning scenarios with students across the engagement and knowledge spectrum.

As AI becomes increasingly proficient, it will soon become ubiquitous, transforming clinical medicine across all healthcare sectors. Investigation of AI has now entered into the era of randomized controlled trials^15^. Additionally, a profusion of pragmatic and observational studies supports a versatile role of AI in virtually all medical disciplines and specialities by improving risk assessment^16,17^, data reduction, clinical decision support^18,19^, operational efficiency, and patient communication^20,21^.

Inspired by the remarkable performance of ChatGPT on the USMLE, clinicians within AnsibleHealth, a virtual chronic pulmonary disease clinic, have begun to experiment with ChatGPT as part of their workflows. Inputting queries in a secure and de-identified manner, our clinicians request ChatGPT to assist with traditionally onerous writing tasks such as composing appeal letters to payors, simplifying radiology reports (and other jargon-dense records) to facilitate patient comprehension, and even to brainstorm freely in a bid to kindle insight when faced with nebulous and diagnostically challenging cases. Overall, our clinicians reported a 33% decrease (future publication) in the time required to complete documentation and indirect patient care tasks. We believe this is an early but important signal that LLMs such as ChatGPT are reaching a maturity level that will soon impact clinical care at large and its ability to deliver truly individualized, compassionate, and scalable healthcare.

## Data Availability

The data analyzed in this study were obtained from USMLE sample questions sets which are publicly available. The question index, raw inputs, and raw AI outputs are available in the Online Data Supplement. Inquiries and requests for additional dataset items and adjudication results can be provided upon reasonable request by contacting Victor Tseng, MD.

## GLOSSARY OF NONSTANDARD ABBREVIATIONS

ACI: Accuracy-Concordance-Insight scoring system
DOI: Density of insight
GPT: Generative pretrained transformer
LLM: Large language model
MCSA: Multiple choice single answer
MC-J: Multiple choice single answer with forced justification
MC-NJ: Multiple choice single answer without forced justification
NLP: Natural language processing
OE: Open-ended question formulation
Q*n*.*m*: Question *n*, input run *m*
USMLE: United States Medical Licensing Exam

## ACKNOWLEDGEMENTS

The authors thank Dr. Kristine Vanijchroenkarn, MD and Ms. Audra Doyle RRT, NP for fruitful discussions and technical assistance. We also thank Mr. Vangjush Vellahu for technical assistance with graphical design and preparation.

## AUTHOR CONTRIBUTIONS

THK, MC, and VT conceived and designed the study, developed the study protocol, supervised the research team, analyzed the data, and wrote the manuscript. AM, CS, LDL, CE, MM, DJC, and JM encoded and input the data into ChatGPT. THK, VT, AM, and CS independently adjudicated the raw ChatGPT outputs. JM and VT performed data synthesis, quality control, and statistical analyses. ChatGPT contributed to the writing of several sections of this manuscript.

---

**Supplemental Table 1.**
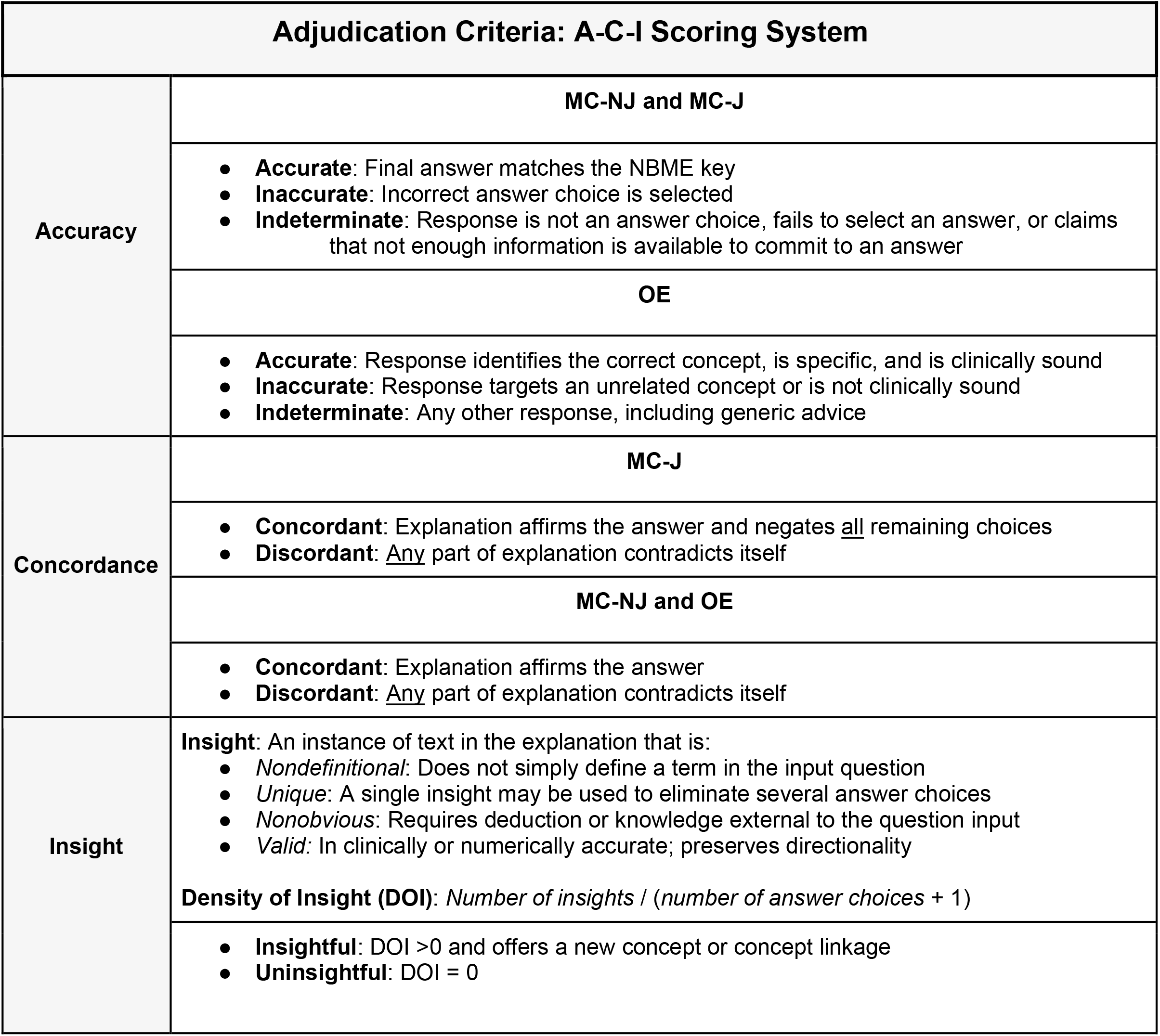

| Adjudication Criteria: A-C-I Scoring System |  |
| --- | --- |
| Accuracy | MC-NJ and MC-J |
|  | • **Accurate:** Final answer matches the NBME key<br>• **Inaccurate:** Incorrect answer choice is selected<br>• **Indeterminate:** Response is not an answer choice, fails to select an answer, or claims that not enough information is available to commit to an answer |
|  | OE |
|  | • **Accurate:** Response identifies the correct concept, is specific, and is clinically sound<br>• **Inaccurate:** Response targets an unrelated concept or is not clinically sound<br>• **Indeterminate:** Any other response, including generic advice |
| Concordance | MC-J |
|  | • **Concordant:** Explanation affirms the answer and negates <u>all</u> remaining choices<br>• **Discordant:** <u>Any</u> part of explanation contradicts itself |
|  | MC-NJ and OE |
|  | • **Concordant:** Explanation affirms the answer<br>• **Discordant:** <u>Any</u> part of explanation contradicts itself |
| Insight | **Insight:** An instance of text in the explanation that is:<br>• *Nondefinitional:* Does not simply define a term in the input question<br>• *Unique:* A single insight may be used to eliminate several answer choices<br>• *Nonobvious:* Requires deduction or knowledge external to the question input<br>• *Valid:* In clinically or numerically accurate; preserves directionality |
| | **Density of Insight (DOI):** $\text{Number of insights} / (\text{number of answer choices} + 1)$ |
|  | • **Insightful:** DOI >0 and offers a new concept or concept linkage<br>• **Uninsightful:** DOI = 0 |

